# Evaluation of an artificial intelligence model for identification of intracranial hemorrhage subtypes on computed tomography of the head

**DOI:** 10.1101/2023.09.07.23295189

**Authors:** James M Hillis, Bernardo C Bizzo, Isabella Newbury-Chaet, Sarah F Mercaldo, John K Chin, Ankita Ghatak, Madeleine A Halle, Eric L’Italien, Ashley L MacDonald, Alex S Schultz, Karen Buch, John Conklin, Stuart Pomerantz, Sandra Rincon, Keith J Dreyer, William A Mehan

## Abstract

**Importance:** Intracranial hemorrhage is a critical finding on computed tomography (CT) of the head.

**Objective:** This study compared the accuracy of an AI model (Annalise Enterprise CTB) to consensus neuroradiologist interpretations in detecting four hemorrhage subtypes: acute subdural/epidural hematoma, acute subarachnoid hemorrhage, intra-axial hemorrhage and intraventricular hemorrhage.

**Design:** A retrospective standalone performance assessment was conducted on datasets of non-contrast CT head cases acquired between 2016 and 2022 for each hemorrhage subtype.

**Setting:** The cases were obtained from five hospitals in the United States.

**Participants:** The cases were obtained from patients aged 18 years or older. The positive cases were selected based on the original clinical reports using natural language processing and manual confirmation. The negative cases were selected by taking the next negative case acquired from the same CT scanner after positive cases.

**Interventions:** Each case was interpreted independently by up to three neuroradiologists to establish consensus interpretations. Each case was then interpreted by the AI model for the presence of the relevant hemorrhage subtype. The neuroradiologists were provided with the entire CT study. The AI model separately received thin (≤1.5mm) and/or thick (>1.5 and ≤5mm) axial series.

**Results:** The four cohorts included 571 cases for acute subdural/epidural hematoma, 310 cases for acute subarachnoid hemorrhage, 926 cases for intra-axial hemorrhage and 199 cases for intraventricular hemorrhage. The AI model identified acute subdural/epidural hematoma with area under the curve (AUC) 0.973 (95% confidence interval (CI), 0.958-0.984) on thin series and 0.942 (95% CI, 0.921-0.959) on thick series; acute subarachnoid hemorrhage with AUC 0.993 (95% CI, 0.984-0.998) on thin series and 0.966 (95% CI, 0.945-0.983) on thick series; intra-axial hemorrhage with AUC 0.969 (95% CI, 0.956-0.980) on thin series and 0.966 (95% CI, 0.953-0.976) on thick series; and intraventricular hemorrhage with AUC 0.987 (95% CI, 0.969-0.997) on thin series and 0.983 (95% CI, 0.968-0.994) on thick series. Each finding had at least one operating point with sensitivity and specificity greater than 80%.

**Conclusions and Relevance:** The assessed AI model accurately identified intracranial hemorrhage subtypes in this CT dataset. Its use could assist the clinical workflow especially through enabling triage of abnormal CTs.

**Key Points:** *Question:* Does a commercial artificial intelligence model accurately identify intracranial hemorrhage subtypes on computed tomography (CT) of the head?

*Findings:* This retrospective study used non-contrast CT studies to compare artificial intelligence model outputs to consensus neuroradiologist interpretations. The model was provided with either thin (≤1.5mm) or thick (>1.5 and ≤5mm) series. The model detected each of acute subdural/epidural hematoma, acute subarachnoid hemorrhage, intra-axial hemorrhage and intraventricular hemorrhage with sensitivity and specificity greater than 80%.

*Meaning:* This artificial intelligence model could assist radiologists through its accurate detection of intracranial hemorrhage subtypes.

## Introduction

Intracranial hemorrhage is a critical finding on computed tomography (CT) of the head and requires emergent medical attention. There are four key subtypes that occur in different regions of the brain: epidural/subdural hematoma, subarachnoid hemorrhage, intra-axial hemorrhage and intraventricular hemorrhage.^1,2^ The faster detection of intracranial hemorrhage on head CT can enable sooner management and intervention including surgery.^3–5^

The use of artificial intelligence (AI) to identify intracranial hemorrhage on head CT has been proposed to assist clinical care, especially by triaging suspected cases for sooner interpretation by a radiologist. There have been at least thirteen computer assisted triage devices (CADt) cleared by the US Food and Drug Administration (FDA) for intracranial hemorrhage detection.^6^ While these devices report subgroup performance for identification of different intracranial hemorrhage subtypes as an “intracranial hemorrhage”,^7,8^ the AI model assessed in the current study is the first FDA-cleared CADt device to output the identified intracranial hemorrhage subtype(s).^9^ It specifically identifies acute subdural/epidural hematoma, acute subarachnoid hemorrhage, intra-axial hemorrhage and intraventricular hemorrhage.

This study calculated the performance of this AI model by comparing its outputs to consensus neuroradiologist interpretations on a cohort of head CT cases for each of the four findings. The AI device was provided separately with thin (≤1.5mm) and/or thick (>1.5 and ≤5mm) axial series from each case so that the performance on different slice thicknesses could be calculated. The performance was also calculated for cases belonging to demographic and technical subgroups to determine the generalizability of the device.

## Methods

### Study design

This retrospective standalone model performance study was conducted using radiology cases from five hospitals within the Mass General Brigham (MGB) network between 2016 and 2022. It was approved by the MGB Institutional Review Board with waiver of informed consent. It was conducted in accordance with relevant guidelines and regulations including the Health Insurance Portability and Accountability Act (HIPAA). This report followed the Standards for Reporting Diagnostic Accuracy (STARD 2015) reporting guideline.

### Case selection

The cohorts for each of the four intracranial hemorrhage subtypes were selected in a consecutive manner based on the original radiology reports. The positive cases were identified through a natural language processing search engine (Nuance mPower Clinical Analytics) followed by manual report review. The negative cases were identified by taking the next negative case acquired on the same CT scanner after the positive cases to avoid temporal and technical bias. The cohort size for each of the positive and negative cases was based on powering calculations as described in the statistical analysis section below. When the cohort had equal numbers of positive and negative cases, the next negative cases were taken after each positive case; when there were unequal numbers of positive and negative cases, the next negative cases were taken after every Nth positive case based on the ratio of positive to negative cases to ensure the principles of consecutive selection applied.

The cohort considered all CT head cases performed at a hospital including inpatient and outpatient; there were no limitations on the original CT head clinical indication. The CT head cases were obtained from patients at least 18 years of age. The CT head cases were taken from unique patients; only the first CT head from a given patient was included. It was possible for a case to be included across multiple cohorts (e.g., both intra-axial hemorrhage and intraventricular hemorrhage).

All cases were deidentified and underwent an image quality review by an American Board of Radiology (ABR)-certified neuroradiologist. The relevant series for the model interpretations were selected at the same time as described under the series selection section below. The review was performed using the FDA-cleared eUnity image visualization software (Version 6 or higher) and an internal web-based annotation system that utilized the REDCap electronic data capture tools hosted at MGB.^10,11^

### Series selection

The model was provided with a single selected series at the time of model inference. These series were non-contrast thin (≤1.5mm) and/or thick (>1.5 and ≤5mm) axial series for each CT head case. The series were selected such that the thin series was the thinnest available series ≤1.5mm; the thick series was randomized between the thinnest and thickest available series >1.5 and ≤5mm to ensure representation of series thicknesses across the entire range. The series were selected at the same time as the image quality review. After series selection, a DICOM metadata review was additionally performed to ensure that the slice thickness was within the appropriate range and that there was a consistent slice interval (with tolerance of ±0.2mm).

### Ground truth interpretations

Ground truth interpretations were performed by up to three ABR-certified neuroradiologists. They answered whether the relevant finding was “Present” or “Absent”. They provided their interpretations independently, without access to the original radiology reports and in different worklist orders. They used the same image visualization software and annotation system as was used in the image quality review. They had access to the entire CT head case (i.e., were not restricted to the series selected for model inference). For determining consensus interpretations, a “2+1” strategy was used: the first two neuroradiologists interpreted every case and a third neuroradiologist then interpreted cases with discrepant interpretations.

### Model inference

The evaluated AI model was version 3.1.0 of the Annalise Enterprise CTB Triage Trauma device. It is the same AI model used by the Annalise Enterprise (CTB module) device, which is commercially available in some non-US markets and whose development has been previously described.^12^ In brief, it consists of an ensemble of five neural networks with three heads: one for classification, one for left-right localization and one for segmentation. It can identify 130 different radiological findings and was trained on over 200,000 CT head cases, which were each labelled by at least three radiologists.

The Annalise Enterprise CTB Triage Trauma device only provides binary classification outputs about the identification of findings, which is consistent with FDA regulations for CADt devices. The model was installed at MGB for use in this study and received only the Digital Imaging and Communications in Medicine (DICOM)-formatted CT head cases. It outputted a classification score between 0 and 1 for each of acute subdural/epidural hematoma, acute subarachnoid hemorrhage, intra-axial hemorrhage and intraventricular hemorrhage. A binary output for each of these findings could be derived using prespecified operating points.

### Statistical analysis

The statistical analysis was performed in R (version 4.0.2) on the full analysis set. The predefined endpoints included the areas under the receiver operating characteristic curves (AUCs) for the identification of acute subdural/epidural hematoma, acute subarachnoid hemorrhage, intra-axial hemorrhage and intraventricular hemorrhage for each of thin and thick series. The AUCs were calculated using the consensus annotations and the classification scores from the AI model. The prespecified endpoints also included the sensitivity and specificity at predetermined operating points; this paper reports the performance at those operating points that have received US Food and Drug Administration clearance. They were calculated by comparing the binary model output at each operating point with the consensus annotations (i.e., by calculating the number of true positive, false negative, true negative and false positive cases).

The AUCs, sensitivities and specificities were calculated as exploratory analyses for the subgroups of sex, age, race, ethnicity and CT scanner manufacturer. These parameters were derived from clinical databases or DICOM fields for each radiology case. Any missing data were treated as “Unknown” and no data were imputed.

All confidence intervals (CIs) were calculated using bootstrapped intervals with 2,000 resamples. The sample sizes for each of the findings were powered based on preliminary model results at a balanced operating point to ensure the lower bound of the 95% CI for sensitivity was >80% and specificity was >80%.

## Results

### Acute subdural/epidural hematoma

A cohort of 571 CT head cases were selected for the acute subdural/epidural hematoma cohort. This cohort resulted in 423 thin series and 571 thick series for which the model could be evaluated (Supplementary Table 1).

#### Thin series

The model successfully performed inference on 409 (96.7%) thin series. This cohort for analysis included 185 (45.2%) women and 224 (54.8%) men; mean (SD) age was 67.0 (19.3) years; there were 308 (75.3%) positive cases and 101 (24.7%) negative cases (Table 1). The AI model identified acute subdural/epidural hematoma with AUC of 0.973 (95% CI: 0.958-0.984; Figure 1A) and, at an operating point of 0.060177, the sensitivity was 91.6% (95% CI: 88.3-94.5%) and the specificity was 87.1% (95% CI: 80.2-93.1%; Supplementary Table 2). These performances were broadly consistent across sex, age, ethnicity, race and manufacturer with all subgroups having sensitivity and specificity of at least 80% except for “unavailable” ethnicity specificity (66.7% for 6 cases) and Asian race specificity (75.0% for 4 cases; Supplementary Table 3).

**Figure 1:**
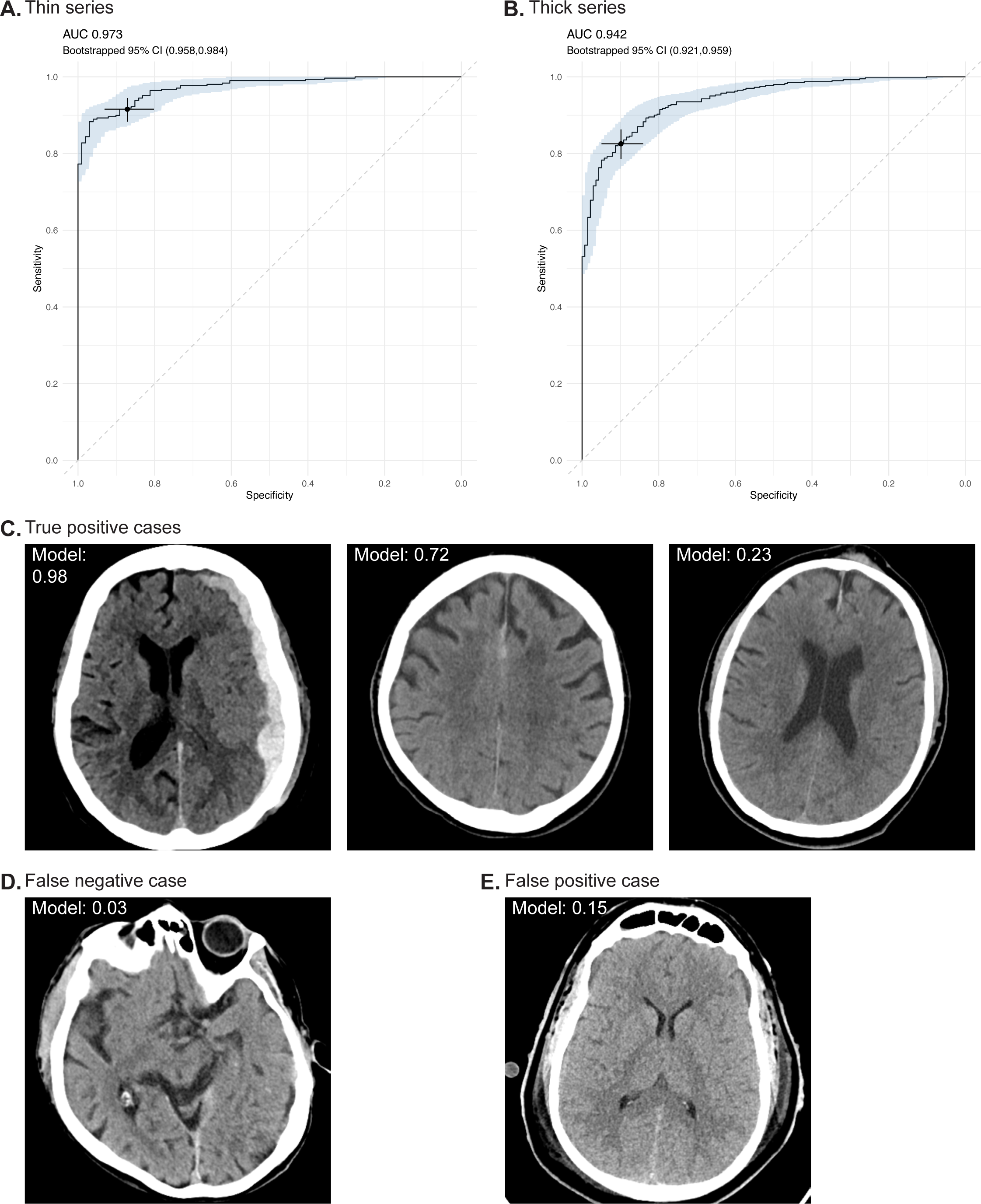
Performance for acute subdural/epidural hematoma. A and B, Receiver operating characteristic curves for the thin series (A) and thick series (B). The shaded region reflects the bootstrapped 95% CIs. The selected point on each graph reflects the operating point at the operating points described in the text. C, D and E, Example images for true positive (C), false negative (D) and false positive (E) cases. The model classification score output is provided for each case.

**Table 1:**
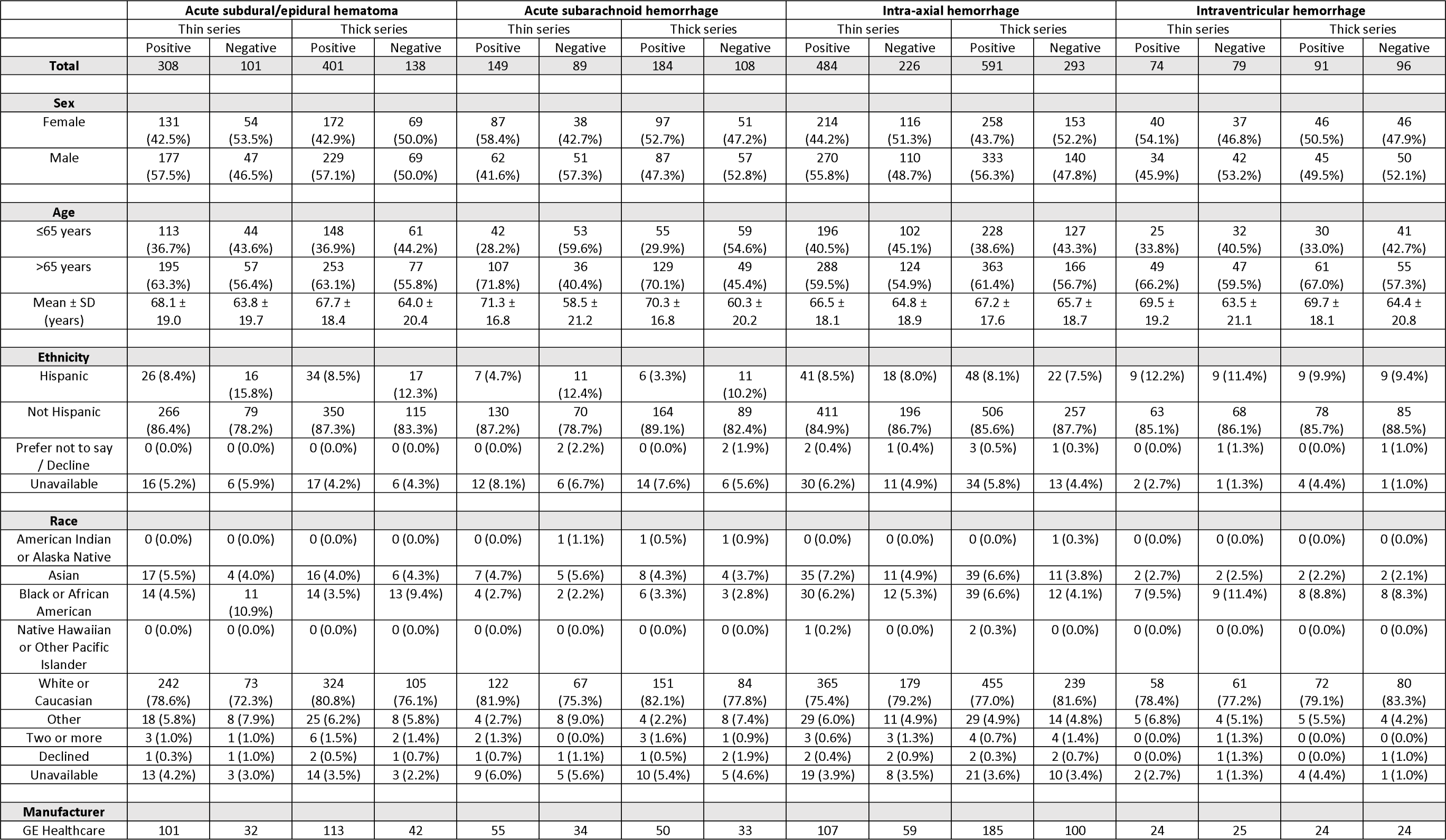
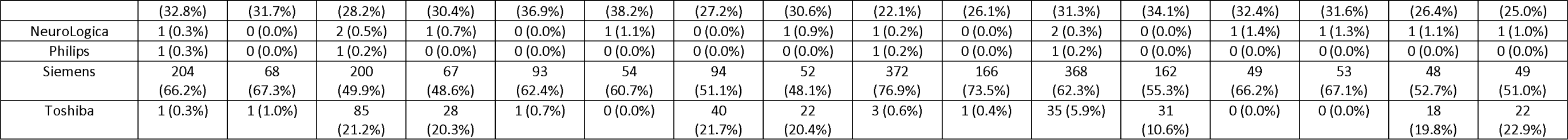
Demographic and technical breakdown of CT head cases for each finding.

#### Thick series

The model successfully performed inference on 539 (94.4%) thick series. This cohort for analysis included 241 (44.7%) women and 298 (55.3%) men; mean (SD) age was 66.8 (19.0) years; there were 401 (74.4%) positive cases and 138 (25.6%) negative cases (Table 1). The AI model identified acute subdural/epidural hematoma with AUC of 0.942 (95% CI: 0.921-0.959; Figure 1B) and, at an operating point of 0.060177, the sensitivity was 82.5% (95% CI: 78.8-86.0%) and the specificity was 89.9% (84.8-94.2%; Supplementary Table 2). These performances were broadly consistent across sex, age, ethnicity, race and manufacturer with all subgroups having sensitivity and specificity of at least 80% except for age ≤65 years sensitivity (77.7% for 148 cases), Black or African American race sensitivity (78.6% for 14 cases), NeuroLogica manufacturer specificity (0.0% for 1 case; Supplementary Table 3).

### Acute subarachnoid hemorrhage

A cohort of 310 CT head cases were selected for the acute subarachnoid hemorrhage cohort. This cohort resulted in 244 thin series and 309 thick series for which the model could be evaluated (Supplementary Table 4).

#### Thin series

The model successfully performed inference on 238 (97.5%) thin series. This cohort for analysis included 125 (52.5%) women and 113 (47.5%) men; mean (SD) age was 66.5 (19.5) years; there were 149 (62.6%) positive cases and 89 (37.4%) negative cases (Table 1). The AI model identified acute subarachnoid hemorrhage with AUC of 0.993 (95% CI: 0.984-0.998; Figure 2A) and, at an operating point of 0.060162, the sensitivity was 94.0% (95% CI: 89.9-97.3%) and the specificity was 96.6% (95% CI: 92.1-100.0%; Supplementary Table 5). These performances were consistent across sex, age, ethnicity, race and manufacturer with all subgroups having sensitivity and specificity of at least 80% (Supplementary Table 6).

**Figure 2:**
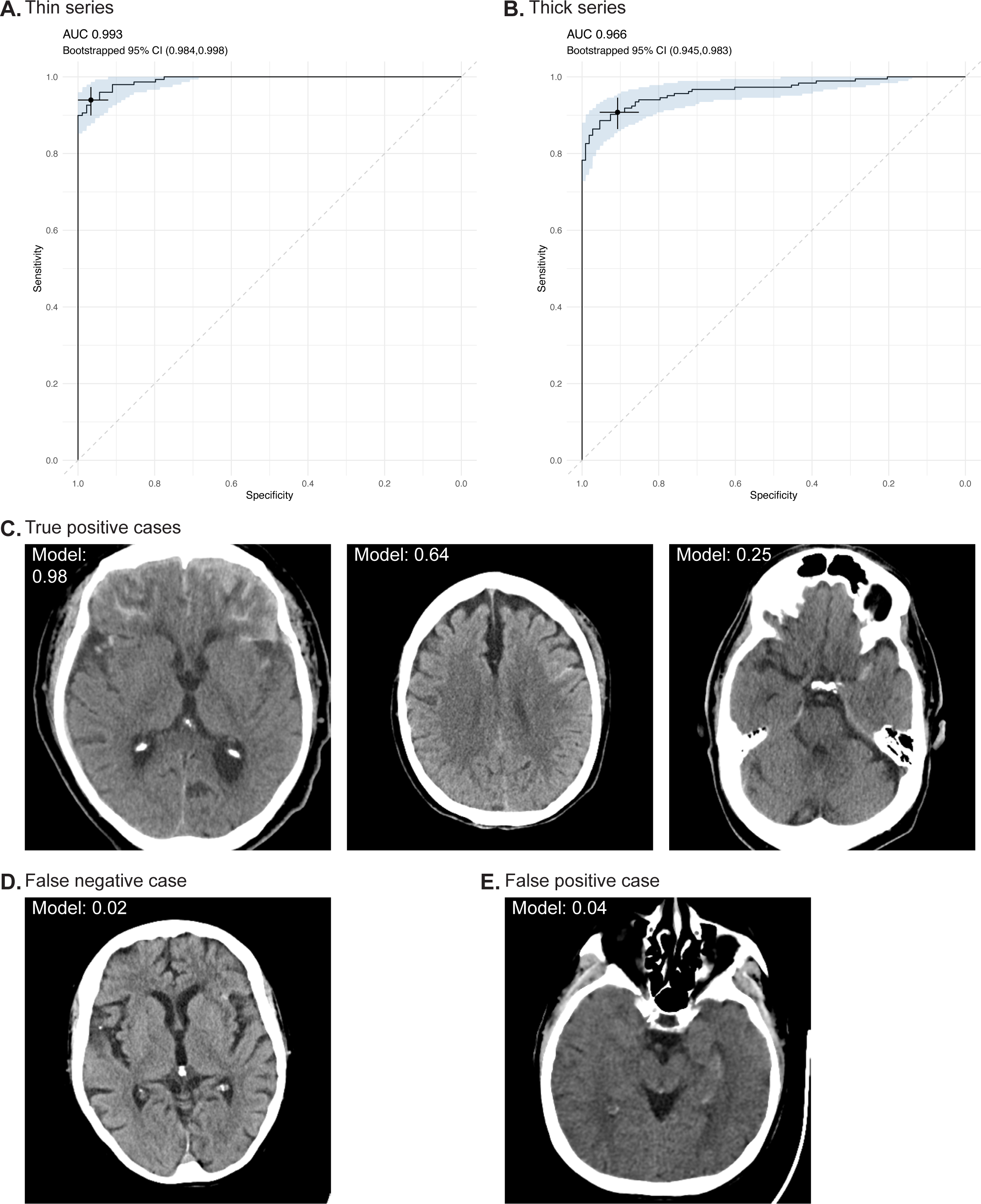
Performance for acute subarachnoid hemorrhage. A and B, Receiver operating characteristic curves for the thin series (A) and thick series (B). The shaded region reflects the bootstrapped 95% CIs. The selected point on each graph reflects the operating point at the operating points described in the text. C, D and E, Example images for true positive (C), false negative (D) and false positive (E) cases. The model classification score output is provided for each case.

#### Thick series

The model successfully performed inference on 292 (94.5%) thick series. This cohort for analysis included 148 (50.7%) women and 144 (49.3%) men; mean (SD) age was 66.6 (18.7) years; there were 184 (63.0%) positive cases and 108 (37.0%) negative cases (Table 1). The AI model identified acute subarachnoid hemorrhage with AUC of 0.966 (95% CI: 0.945-0.983; Figure 2B) and, at an operating point of 0.020255, the sensitivity was 90.8% (95% CI: 86.4-94.6%) and the specificity was 90.7% (95% CI: 85.2-95.4%; Supplementary Table 5). These performances were consistent across sex, age, ethnicity, race and manufacturer with all subgroups having sensitivity and specificity of at least 80% (Supplementary Table 6).

### Intra-axial hemorrhage

A cohort of 926 CT head cases were selected for the intra-axial hemorrhage cohort. This cohort resulted in 733 thin series and 925 thick series for which the model could be evaluated (Supplementary Table 7).

#### Thin series

The model successfully performed inference on 710 (96.9%) thin series. This cohort for analysis included 330 (46.5%) women and 380 (53.5%) men; mean (SD) age was 66.0 (18.4) years; there were 484 (68.2%) positive cases and 226 (31.8%) negative cases (Table 1). The AI model identified intra-axial hemorrhage with AUC of 0.969 (95% CI: 0.956-0.980; Figure 3A) and, at an operating point of 0.322700, the sensitivity was 93.2% (95% CI: 90.9-95.5%) and the specificity was 85.8% (95% CI: 81.0-90.3%; Supplementary Table 8). These performances were broadly consistent across sex, age, ethnicity, race and manufacturer with all subgroups having sensitivity and specificity of at least 80% except for “two or more” race sensitivity (66.7% for 3 cases), “unavailable” ethnicity specificity (54.5% for 11 cases), Asian race specificity (72.7% for 11 cases) and “unavailable” race specificity (75.0% for 8 cases; Supplementary Table 9).

**Figure 3:**
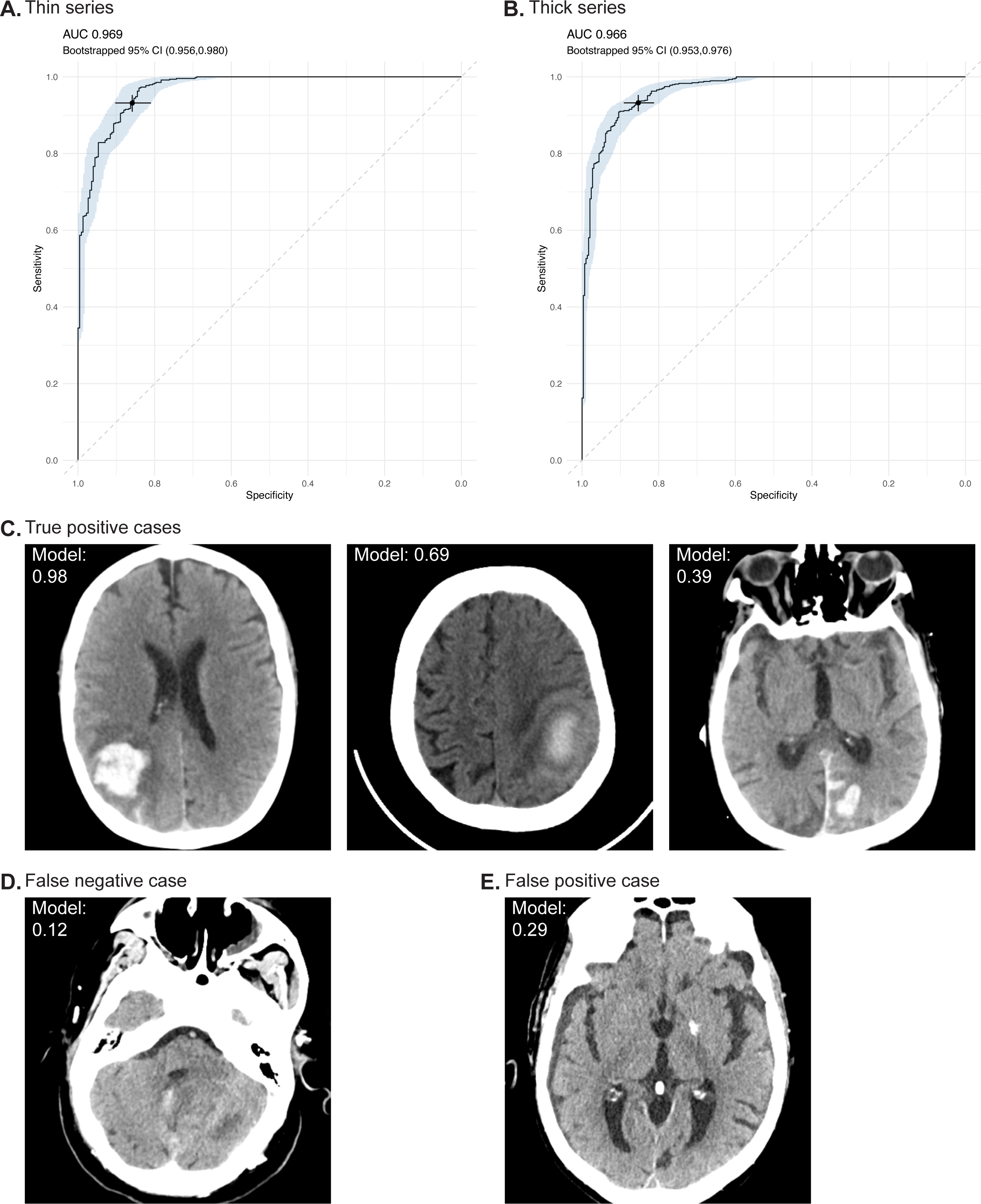
Performance for intra-axial hemorrhage. A and B, Receiver operating characteristic curves for the thin series (A) and thick series (B). The shaded region reflects the bootstrapped 95% CIs. The selected point on each graph reflects the operating point at the operating points described in the text. C, D and E, Example images for true positive (C), false negative (D) and false positive (E) cases. The model classification score output is provided for each case.

#### Thick series

The model successfully performed inference on 884 (95.6%) thick series. This cohort for analysis included 411 (46.5%) women and 473 (53.5%) men; mean (SD) age was 66.7 (18.0) years; there were 591 (66.9%) positive cases and 293 (33.1%) negative cases (Table 1). The AI model identified intra-axial hemorrhage with AUC of 0.966 (95% CI: 0.953-0.976; Figure 3A) and, at an operating point of 0.203600, the sensitivity was 93.2% (95% CI: 91.2-95.3%) and the specificity was 85.3% (95% CI: 80.9-89.1%; Supplementary Table 8). These performances were broadly consistent across sex, age, ethnicity, race and manufacturer with all subgroups having sensitivity and specificity of at least 80% except for “two or more” race sensitivity (75.0% for 4 cases), NeuroLogica manufacturer sensitivity (50.0% for 2 cases), “unavailable” ethnicity specificity (46.2% for 13 cases), Asian race specificity (72.7% for 11 cases) and “unavailable” race specificity (50.0% for 10 cases; Supplementary Table 9).

### Intraventricular hemorrhage

A cohort of 199 CT head cases were selected for the intraventricular hemorrhage cohort. This cohort resulted in 159 thin series and 199 thick series for which the model could be evaluated (Supplementary Table 10).

#### Thin series

The model successfully performed inference on 153 (96.2%) thin series. This cohort for analysis included 77 (50.3%) women and 76 (49.7%) men; mean (SD) age was 66.4 (20.4) years; there were 74 (48.4%) positive cases and 79 (51.6%) negative cases (Table 1). The AI model identified intraventricular hemorrhage with AUC of 0.987 (95% CI: 0.969-0.997; Figure 4A) and, at an operating point of 0.051859, the sensitivity was 90.5% (95% CI: 83.8-95.9%) and the specificity was 97.5% (95% CI: 93.7-100.0%; Supplementary Table 11). These performances were broadly consistent across sex, age, ethnicity, race and manufacturer with all subgroups having sensitivity and specificity of at least 80% except for NeuoLogica sensitivity (0.0% for 1 case; Supplementary Table 12).

**Figure 4:**
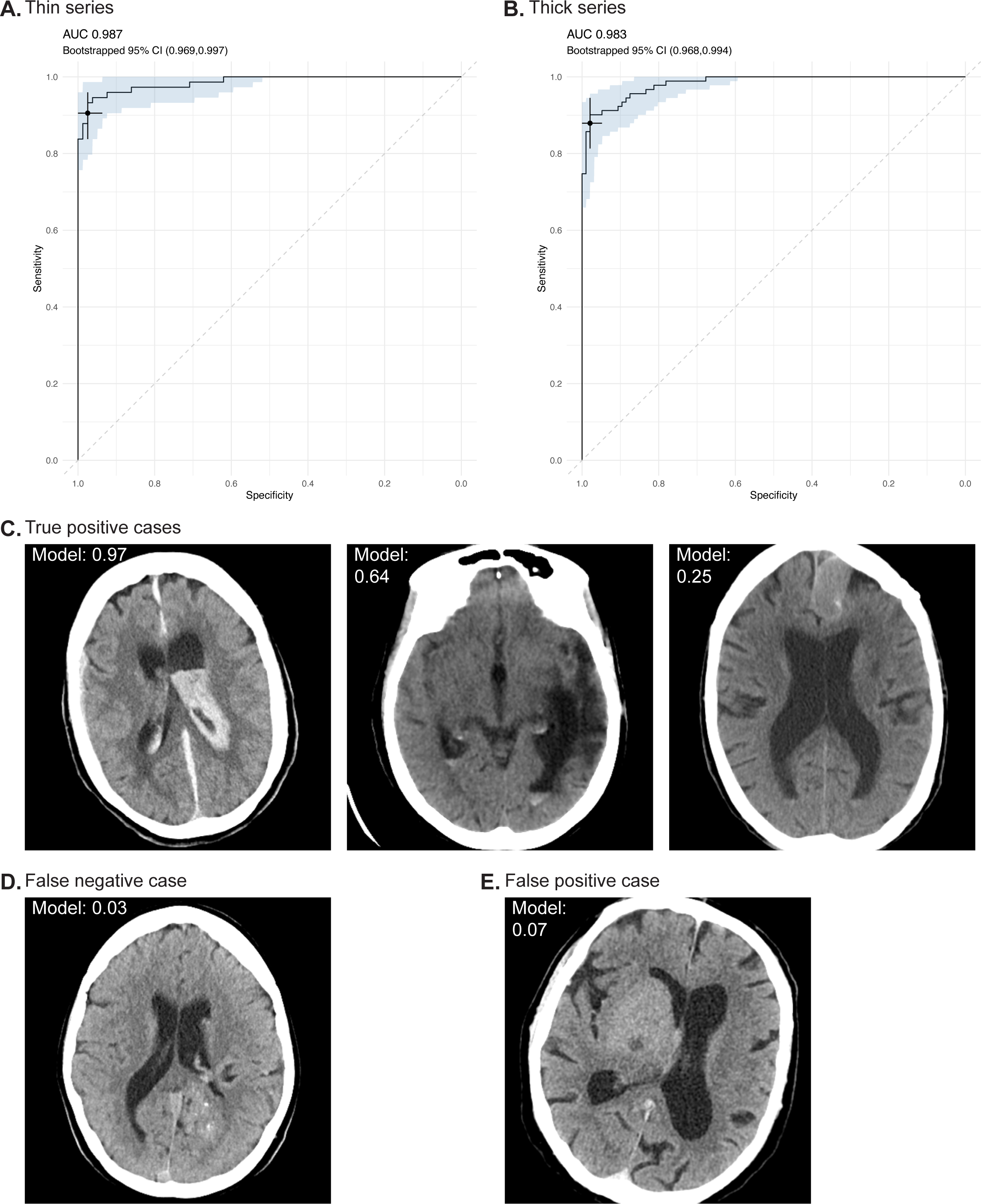
Performance for intraventricular hemorrhage. A and B, Receiver operating characteristic curves for the thin series (A) and thick series (B). The shaded region reflects the bootstrapped 95% CIs. The selected point on each graph reflects the operating point at the operating points described in the text. C, D and E, Example images for true positive (C), false negative (D) and false positive (E) cases. The model classification score output is provided for each case.

#### Thick series

The model successfully performed inference on 187 (94.0%) thick series. This cohort for analysis included 92 (49.2%) women and 95 (50.8%) men; mean (SD) age was 67.0 (19.6) years; there were 91 (48.7%) positive cases and 96 (51.3%) negative cases (Table 1). The AI model identified intraventricular hemorrhage with AUC of 0.983 (95% CI: 0.968-0.994; Figure 4A) and, at an operating point of 0.051859, the sensitivity was 87.9% (95% CI: 81.3-93.4%) and the specificity was 97.9% (95% CI: 94.8-100.0%; Supplementary Table 11). These performances were consistent across sex, age, ethnicity, race and manufacturer with all subgroups having sensitivity and specificity of at least 80% (Supplementary Table 12).

## Discussion

This retrospective diagnostic study assessed the performance of an AI model in identifying acute subdural/epidural hematoma, acute subarachnoid hemorrhage, intra-axial hemorrhage and intraventricular hemorrhage on head CT. The model achieved sensitivity greater than 80% and specificity greater than 80% for all four findings. These results are consistent with other FDA-cleared intracranial hemorrhage CADt devices.^7,8,13–22^ However, this model is the first FDA-cleared CADt device that outputs the subtype of intracranial hemorrhage.^9^

The output of the subtype of intracranial hemorrhage provides users with more explainability of the model outputs. As has been noted on an assessment of a similar model for pneumothorax identification, the FDA regulations for a CADt device only permit devices to output the binary classification performance (the AI model assessed here technically outputs a binary classification for each of the four subtypes).^23^ A segmentation output could otherwise further assist with explainability by demonstrating the location of the identified intracranial hemorrhage.

The performance of the model was broadly consistent across sex, age, ethnicity, race and manufacturer subgroups with the vast majority achieving sensitivity and specificity above 80%. For the minority of subgroups that did not achieve a sensitivity or specificity of at least 80%, all except one had small sample sizes of less than 15. The sensitivity for the age ≤65 years subgroup for the thick series for acute subdural/epidural hematoma was 77.7% despite including 148 cases; we note that the overall sensitivity was 82.5% and that these two values had overlapping 95% CIs.

When the model performance was calculated at the same operating point on both thin and thick series, it had better sensitivity on the thin series and better specificity on the thick series. This observation likely relates to the increased z-axis resolution on the thin series, which can facilitate better identification of hemorrhage especially for small hemorrhages that may only be present on a small number of slices. The thinner series, however, also have more noise that could lead to the model incorrectly identifying hemorrhage.^24^ A consideration for clinical use may be the use of a lower operating point for thick series to balance out this difference.

As part of the study design, the cohorts of cases were selected based on the original radiology reports and subsequently interpreted in a consensus manner by neuroradiologists. The rationale for this approach is that the binary presence or absence of each finding is confirmed for every case based on the radiologic images alone, which matches what the model uses. It is, however, possible for the interpretations to change between the original reports and the neuroradiologist interpretations especially given the original reports can be aided by knowledge of the clinical situation and longitudinal radiology scans. All cohorts had a smaller number of positive cases based on the consensus neuroradiologist interpretations compared to the original reports. These cases would be considered ground truth negative for the analysis and, if the model identified them as positive, then they would have been recorded as false positive cases with subsequently decreased specificity. The real-world specificity may therefore be higher than what is recorded as part of this study.

### Limitations

A key limitation of this study is that it is a retrospective study outside of the clinical workflow. As was noted in the similar assessment of a pneumothorax model, this study therefore establishes the accuracy of the model but does not assess its impact on the clinical workflow including for case prioritization and patient outcomes.^23^ This initial step is necessary to ensure the model has the potential to provide clinical benefit. Further evaluation will be required moving forward to prove such benefit.

This study examines the ability of the model to identify each hemorrhage subtype independently. It does not consider the overlap of hemorrhage subtypes, which is common in the clinical environment; as an example, intraventricular hemorrhage occurs in 30 to 50% of patients with intra-axial hemorrhage.^5^ This overlap may lead to redundancy especially when the model identifies a first hemorrhage subtype but then misses a second or third subtype. It is also possible that the model incorrectly identifies a second or third subtype when only a first subtype is present. Further research may consider the application of this model when multiple hemorrhage subtypes are present.

## Conclusion

This diagnostic study assessed an AI model that accurately detected four subtypes of intracranial hemorrhage including acute subdural/epidural hematoma, acute subarachnoid hemorrhage, intra-axial hemorrhage and intraventricular hemorrhage. Its use in the clinical environment may lead to improved care and outcomes for patients with intracranial hemorrhage.

## Supporting information

STARD Checklist

## Data Availability

The AI model is part of the FDA cleared Annalise Enterprise CTB Triage Trauma device. The test dataset generated for this study contains protected patient information. Some data may be available for research purposes from the corresponding author upon reasonable request.

## Acknowledgements

The authors thank the broader Mass General Brigham Data Science Office and Annalise teams for their assistance with this project.

## Access to data and data analysis

JMH had full access to all the data in the study and takes responsibility for the integrity of the data and the accuracy of the data analysis. SFM performed the statistical analyses.

