## Supplementary Tables 1-12 for "Evaluation of an artificial intelligence model for identification of intracranial hemorrhage subtypes on computed tomography of the head"

**Supplementary Table 1**: Number of cases for acute subdural/epidural hematoma cohort. *The number of selected cases was based on powering calculations; the ground truth radiologists did not have access to the original radiology reports and the interpretation could change.

|  | Positive cases | Negative cases | Total cases |
| --- | --- | --- | --- |
| **Original cases** |  |  |  |
| Number of cases selected based on original radiology reports* | 471 | 100 | 571 |
| Number of cases following ground truth radiologist interpretations | 428 | 143 | 571 |
| **Thin series (≤1.5mm)** |  |  |  |
| Number of thin series | 316 | 107 | 423 |
| Number of thin series with successful model inference | 308 | 101 | 409 |
| **Thick series (>1.5mm and ≤5mm)** |  |  |  |
| Number of thick series | 428 | 143 | 571 |
| Number of thick series with successful model inference | 401 | 138 | 539 |

**Supplementary Table 2**: Model performance for identification of acute subdural/epidural hematoma at prespecified operating points.

| Operating point | True positive | False negative | Sensitivity (95% CI) | True negative | False positive | Specificity (95% CI) |
| --- | --- | --- | --- | --- | --- | --- |
| **Thin series** |  |  |  |  |  |  |
| 0.060177 | 282 | 26 | 91.6 (88.3-94.5) | 88 | 13 | 87.1 (80.2-93.1) |
| 0.101143 | 275 | 33 | 89.3 (85.7-92.5) | 96 | 5 | 95.0 (90.1-99.0) |
| 0.135700 | 267 | 41 | 86.7 (82.8-90.3) | 98 | 3 | 97.0 (93.1-100.0) |
| **Thick series** |  |  |  |  |  |  |
| 0.060177 | 331 | 70 | 82.5 (78.8-86.0) | 124 | 14 | 89.9 (84.8-94.2) |

**Supplementary Table 3**: Demographic and technical subgroup performance for identifying acute subdural/epidural hematoma.

|  | **Thin Series (Operating Point 0.060177)** | | | | | **Thick Series (Operating Point 0.060177)** | | | | |
| --- | --- | --- | --- | --- | --- | --- | --- | --- | --- | --- |
|  | Positive N | Negative N | AUC (95% CI) | Sensitivity (95% CI) | Specificity (95% CI) | Positive N | Negative N | AUC (95% CI) | Sensitivity (95% CI) | Specificity (95% CI) |
| **Overall** | 308 | 101 | 0.973 (0.958-0.984) | 91.6 (88.3-94.5) | 87.1 (80.2-93.1) | 401 | 138 | 0.942 (0.921-0.959) | 82.5 (78.8-86.0) | 89.9 (84.8-94.2) |
| **Sex** |  |  |  |  |  |  |  |  |  |  |
| Female | 131 | 54 | 0.962 (0.934-0.983) | 88.5 (82.4-93.9) | 87.0 (77.8-96.3) | 172 | 69 | 0.935 (0.900-0.961) | 80.2 (73.8-86.0) | 91.3 (84.1-97.1) |
| Male | 177 | 47 | 0.983 (0.968-0.993) | 93.8 (90.4-97.2) | 87.2 (76.6-95.7) | 229 | 69 | 0.947 (0.919-0.970) | 84.3 (79.5-88.6) | 88.4 (79.7-95.7) |
| **Age** |  |  |  |  |  |  |  |  |  |  |
| ≤65 years | 113 | 44 | 0.982 (0.961-0.994) | 91.2 (85.8-95.6) | 93.2 (84.1-100.0) | 148 | 61 | 0.945 (0.912-0.970) | 77.7 (70.9-83.8) | 93.4 (86.9-98.4) |
| >65 years | 195 | 57 | 0.970 (0.950-0.985) | 91.8 (87.7-95.4) | 82.5 (71.9-91.2) | 253 | 77 | 0.939 (0.910-0.964) | 85.4 (81.0-89.3) | 87.0 (77.9-93.5) |
| **Ethnicity** |  |  |  |  |  |  |  |  |  |  |
| Hispanic | 26 | 16 | 1.000 (1.000-1.000) | 92.3 (80.8-100.0) | 100.0 (100.0-100.0) | 34 | 17 | 0.990 (0.955-1.000) | 82.4 (67.6-94.1) | 100.0 (100.0-100.0) |
| Not Hispanic | 266 | 79 | 0.970 (0.952-0.983) | 91.0 (87.6-94.4) | 86.1 (78.5-93.7) | 350 | 115 | 0.936 (0.913-0.958) | 82.0 (78.0-86.0) | 88.7 (82.6-93.9) |
| Prefer not to say / Decline | 0 | 0 | - | - | - | 0 | 0 | - | - | - |
| Unavailable | 16 | 6 | 1.000 (1.000-1.000) | 100.0 (100.0-100.0) | 66.7 (33.3-100.0) | 17 | 6 | 0.990 (0.912-1.000) | 94.1 (82.4-100.0) | 83.3 (50.0-100.0) |
| **Race** |  |  |  |  |  |  |  |  |  |  |
| American Indian or Alaska Native | 0 | 0 | - | - | - | 0 | 0 | - | - | - |
| Asian | 17 | 4 | 0.971 (0.838-1.000) | 94.1 (82.4-100.0) | 75.0 (25.0-100.0) | 16 | 6 | 0.938 (0.792-1.000) | 81.2 (62.5-100.0) | 83.3 (50.0-100.0) |
| Black or African American | 14 | 11 | 1.000 (0.961-1.000) | 100.0 (100.0-100.0) | 81.8 (54.5-100.0) | 14 | 13 | 0.984 (0.923-1.000) | 78.6 (57.1-100.0) | 92.3 (76.9-100.0) |
| Native Hawaiian or Other Pacific Islander | 0 | 0 | - | - | - | 0 | 0 | - | - | - |
| White or Caucasian | 242 | 73 | 0.971 (0.953-0.984) | 90.9 (87.2-94.2) | 86.3 (78.1-93.2) | 324 | 105 | 0.935 (0.908-0.956) | 81.8 (77.8-85.8) | 88.6 (81.9-94.3) |
| Other | 18 | 8 | 1.000 (1.000-1.000) | 83.3 (66.7-100.0) | 100.0 (100.0-100.0) | 25 | 8 | 0.975 (0.895-1.000) | 80.0 (64.0-92.0) | 100.0 (100.0-100.0) |
| Two or more | 3 | 1 | 1.000 (1.000-1.000) | 100.0 (100.0-100.0) | 100.0 (100.0-100.0) | 6 | 2 | 1.000 (1.000-1.000) | 100.0 (100.0-100.0) | 100.0 (100.0-100.0) |
| Declined | 1 | 1 | 1.000 (1.000-1.000) | 100.0 (100.0-100.0) | 100.0 (100.0-100.0) | 2 | 1 | 1.000 (1.000-1.000) | 100.0 (100.0-100.0) | 100.0 (100.0-100.0) |
| Unavailable | 13 | 3 | 1.000 (1.000-1.000) | 100.0 (100.0-100.0) | 100.0 (100.0-100.0) | 14 | 3 | 1.000 (1.000-1.000) | 92.9 (78.6-100.0) | 100.0 (100.0-100.0) |
| **Manufacturer** |  |  |  |  |  |  |  |  |  |  |
| GE Healthcare | 101 | 32 | 0.989 (0.973-0.998) | 95.0 (91.1-99.0) | 87.5 (75.0-96.9) | 113 | 42 | 0.964 (0.934- 0.983) | 85.0 (78.7-91.2) | 90.5 (81.0-97.6) |
| NeuroLogica | 1 | 0 | - | 100.0 (100.0-100.0) | - | 2 | 1 | 1.000 (1.000- 1.000) | 100.0 (100.0-100.0) | 0.0 (0.0-0.0) |
| Philips | 1 | 0 | - | 100.0 (100.0-100.0) | - | 1 | 0 | - | 100.0 (100.0-100.0) | - |
| Siemens | 204 | 68 | 0.964 (0.941-0.981) | 89.7 (85.3-93.6) | 86.8 (77.9-94.1) | 200 | 67 | 0.941 (0.911- 0.965) | 81.5 (76.0-86.5) | 94.0 (88.1-98.5) |
| Toshiba | 1 | 1 | 1.000 (1.000-1.000) | 100.0 (100.0-100.0) | 100.0 (100.0-100.0) | 85 | 28 | 0.934 (0.874- 0.974) | 81.2 (72.9-89.4) | 82.1 (67.9-96.4) |

**Supplementary Table 4**: Number of cases for acute subarachnoid hemorrhage cohort. *The number of selected cases was based on powering calculations; the ground truth radiologists did not have access to the original radiology reports and the interpretation could change.

|  | Positive cases | Negative cases | Total cases |
| --- | --- | --- | --- |
| **Original cases** |  |  |  |
| Number of cases selected based on original radiology reports* | 210 | 100 | 310 |
| Number of cases following ground truth radiologist interpretations | 196 | 114 | 310 |
| **Thin series (≤1.5mm)** |  |  |  |
| Number of thin series | 154 | 90 | 244 |
| Number of thin series with successful model inference | 149 | 89 | 238 |
| **Thick series (>1.5mm and ≤5mm)** |  |  |  |
| Number of thick series | 196 | 113 | 309 |
| Number of thick series with successful model inference | 184 | 108 | 292 |

**Supplementary Table 5**: Model performance for identification of acute subarachnoid hemorrhage at prespecified operating points.

| Operating point | True positive | False negative | Sensitivity (95% CI) | True negative | False positive | Specificity (95% CI) |
| --- | --- | --- | --- | --- | --- | --- |
| **Thin series** |  |  |  |  |  |  |
| 0.014372 | 146 | 3 | 98.0 (95.3-100.0) | 80 | 9 | 89.9 (83.1-95.5) |
| 0.060162 | 140 | 9 | 94.0 (89.9-97.3) | 86 | 3 | 96.6 (92.1-100.0) |
| 0.082652 | 134 | 15 | 89.9 (84.6-94.6) | 89 | 0 | 100.0 (100.0-100.0) |
| **Thick series** |  |  |  |  |  |  |
| 0.020255 | 167 | 17 | 90.8 (86.4-94.6) | 98 | 10 | 90.7 (85.2-95.4) |
| 0.030010 | 161 | 23 | 87.5 (82.6-91.8) | 103 | 5 | 95.4 (90.7-99.1) |

**Supplementary Table 6**: Demographic and technical subgroup performance for identifying acute subarachnoid hemorrhage.

|  | **Thin Series (Operating Point 0.060162)** | | | | | **Thick Series (Operating Point 0.020255)** | | | | |
| --- | --- | --- | --- | --- | --- | --- | --- | --- | --- | --- |
|  | Positive N | Negative N | AUC (95% CI) | Sensitivity (95% CI) | Specificity (95% CI) | Positive N | Negative N | AUC (95% CI) | Sensitivity (95% CI) | Specificity (95% CI) |
| **Overall** | 149 | 89 | 0.993 (0.984-0.998) | 94.0 (89.9-97.3) | 96.6 (92.1-100.0) | 184 | 108 | 0.966 (0.945-0.983) | 90.8 (86.4-94.6) | 90.7 (85.2-95.4) |
| **Sex** |  |  |  |  |  |  |  |  |  |  |
| Female | 87 | 38 | 0.992 (0.976-0.998) | 93.1 (87.4-97.7) | 97.4 (92.1-100.0) | 97 | 51 | 0.964 (0.935-0.986) | 88.7 (81.4-94.8) | 92.2 (84.3-98.0) |
| Male | 62 | 51 | 0.996 (0.987-1.000) | 95.2 (90.3-100.0) | 96.1 (90.2-100.0) | 87 | 57 | 0.970 (0.936-0.992) | 93.1 (87.4-97.7) | 89.5 (80.7-96.5) |
| **Age** |  |  |  |  |  |  |  |  |  |  |
| ≤65 years | 42 | 53 | 0.996 (0.984-1.000) | 95.2 (88.1-100.0) | 94.3 (86.8-100.0) | 55 | 59 | 0.970 (0.931-0.995) | 94.5 (87.3-100.0) | 88.1 (79.7-96.6) |
| >65 years | 107 | 36 | 0.995 (0.987-1.000) | 93.5 (88.8-97.2) | 100.0 (100.0-100.0) | 129 | 49 | 0.969 (0.944-0.988) | 89.1 (83.7-94.6) | 93.9 (85.7-100.0) |
| **Ethnicity** |  |  |  |  |  |  |  |  |  |  |
| Hispanic | 7 | 11 | 1.000 (0.922-1.000) | 85.7 (57.1-100.0) | 90.9 (72.7-100.0) | 6 | 11 | 1.000 (1.000-1.000) | 100.0 (100.0-100.0) | 100.0 (100.0-100.0) |
| Not Hispanic | 130 | 70 | 0.993 (0.984-0.998) | 94.6 (90.8-98.5) | 97.1 (92.9-100.0) | 164 | 89 | 0.965 (0.941-0.982) | 90.9 (86.0-95.1) | 89.9 (83.1-95.5) |
| Prefer not to say / Decline | 0 | 2 | - | - | 100.0 (100.0-100.0) | 0 | 2 | - | - | 100.0 (100.0-100.0) |
| Unavailable | 12 | 6 | 1.000 (0.917-1.000) | 91.7 (75.0-100.0) | 100.0 (100.0-100.0) | 14 | 6 | 0.976 (0.881-1.000) | 85.7 (64.3-100.0) | 83.3 (50.0-100.0) |
| **Race** |  |  |  |  |  |  |  |  |  |  |
| American Indian or Alaska Native | 0 | 1 | - | - | 100.0 (100.0-100.0) | 1 | 1 | 1.000 (1.000-1.000) | 100.0 (100.0-100.0) | 100.0 (100.0-100.0) |
| Asian | 7 | 5 | 1.000 (1.000-1.000) | 100.0 (100.0-100.0) | 100.0 (100.0-100.0) | 8 | 4 | 1.000 (1.000-1.000) | 100.0 (100.0-100.0) | 100.0 (100.0-100.0) |
| Black or African American | 4 | 2 | 1.000 (1.000-1.000) | 100.0 (100.0-100.0) | 100.0 (100.0-100.0) | 6 | 3 | 1.000 (1.000-1.000) | 100.0 (100.0-100.0) | 100.0 (100.0-100.0) |
| Native Hawaiian or Other Pacific Islander | 0 | 0 | - | - | - | 0 | 0 | - | - | - |
| White or Caucasian | 122 | 67 | 0.993 (0.984-0.998) | 93.4 (89.3-97.5) | 97.0 (92.5-100.0) | 151 | 84 | 0.959 (0.932-0.978) | 89.4 (84.1-94.0) | 89.3 (82.1-95.2) |
| Other | 4 | 8 | 1.000 (1.000-1.000) | 100.0 (100.0-100.0) | 87.5 (62.5-100.0) | 4 | 8 | 1.000 (1.000-1.000) | 100.0 (100.0-100.0) | 100.0 (100.0-100.0) |
| Two or more | 2 | 0 | - | 100.0 (100.0-100.0) | - | 3 | 1 | 1.000 (1.000-1.000) | 100.0 (100.0-100.0) | 100.0 (100.0-100.0) |
| Declined | 1 | 1 | 1.000 (1.000-1.000) | 100.0 (100.0-100.0) | 100.0 (100.0-100.0) | 1 | 2 | 1.000 (1.000-1.000) | 100.0 (100.0-100.0) | 100.0 (100.0-100.0) |
| Unavailable | 9 | 5 | 1.000 (0.867-1.000) | 88.9 (66.7-100.0) | 100.0 (100.0-100.0) | 10 | 5 | 1.000 (0.880-1.000) | 90.0 (70.0-100.0) | 80.0 (40.0-100.0) |
| **Manufacturer** |  |  |  |  |  |  |  |  |  |  |
| GE Healthcare | 55 | 34 | 0.997 (0.987-1.000) | 94.5 (87.3-100.0) | 94.1 (85.3-100.0) | 50 | 33 | 0.977 (0.942-0.996) | 92.0 (84.0-98.0) | 90.9 (81.8-100.0) |
| NeuroLogica | 0 | 1 | - | - | 100.0 (100.0-100.0) | 0 | 1 | - | - | 100.0 (100.0-100.0) |
| Philips | 0 | 0 | - | - | - | 0 | 0 | - | - | - |
| Siemens | 93 | 54 | 0.990 (0.976-0.998) | 93.5 (88.2-97.8) | 98.1 (94.4-100.0) | 94 | 52 | 0.954 (0.919-0.982) | 88.3 (81.9-94.7) | 88.5 (78.8-96.2) |
| Toshiba | 1 | 0 | - | 100.0 (100.0-100.0) | - | 40 | 22 | 0.984 (0.945-1.000) | 95.0 (87.5-100.0) | 95.5 (86.4-100.0) |

**Supplementary Table 7**: Number of cases for intra-axial hemorrhage cohort. *The number of selected cases was based on powering calculations; the ground truth radiologists did not have access to the original radiology reports and the interpretation could change. **One case was incorrectly processed during the series selection, image quality review and DICOM review when it should have been excluded; its later removal resulted in the cohort having one fewer case than initially planned and is reflected from the second row of this table onwards (which has one fewer case than the first row).

|  | Positive cases | Negative cases | Total cases |
| --- | --- | --- | --- |
| **Original cases** |  |  |  |
| Number of cases selected based on original radiology reports* | 717 | 210 | 927 |
| Number of cases following ground truth radiologist interpretations** | 617 | 309 | 926 |
| **Thin series (≤1.5mm)** |  |  |  |
| Number of thin series | 503 | 230 | 733 |
| Number of thin series with successful model inference | 484 | 226 | 710 |
| **Thick series (>1.5mm and ≤5mm)** |  |  |  |
| Number of thick series | 617 | 308 | 925 |
| Number of thick series with successful model inference | 591 | 293 | 884 |

**Supplementary Table 8**: Model performance identification of intra-axial hemorrhage at prespecified operating points.

| Operating point | True positive | False negative | Sensitivity (95% CI) | True negative | False positive | Specificity (95% CI) |
| --- | --- | --- | --- | --- | --- | --- |
| **Thin series** |  |  |  |  |  |  |
| 0.322700 | 451 | 33 | 93.2 (90.9-95.5) | 194 | 32 | 85.8 (81.0-90.3) |
| **Thick series** |  |  |  |  |  |  |
| 0.203600 | 551 | 40 | 93.2 (91.2-95.3) | 250 | 43 | 85.3 (80.9-89.1) |
| 0.322700 | 533 | 58 | 90.2 (87.6-92.6) | 265 | 28 | 90.4 (87.0-93.9) |

**Supplementary Table 9**: Demographic and technical subgroup performance for identifying intra-axial hemorrhage.

|  | **Thin Series (Operating Point 0.322700)** | | | | | **Thick Series (Operating Point 0.203600)** | | | | |
| --- | --- | --- | --- | --- | --- | --- | --- | --- | --- | --- |
|  | Positive N | Negative N | AUC (95% CI) | Sensitivity (95% CI) | Specificity (95% CI) | Positive N | Negative N | AUC (95% CI) | Sensitivity (95% CI) | Specificity (95% CI) |
| **Overall** | 484 | 226 | 0.969 (0.956-0.980) | 93.2 (90.9-95.5) | 85.8 (81.0-90.3) | 591 | 293 | 0.966 (0.953-0.976) | 93.2 (91.2-95.3) | 85.3 (80.9-89.1) |
| **Sex** |  |  |  |  |  |  |  |  |  |  |
| Female | 214 | 116 | 0.974 (0.958-0.987) | 92.5 (88.8-95.8) | 88.8 (82.8-94.0) | 258 | 153 | 0.968 (0.951-0.981) | 93.0 (89.5-96.1) | 86.3 (81.0-91.5) |
| Male | 270 | 110 | 0.963 (0.939-0.980) | 93.7 (90.7-96.7) | 82.7 (75.5-89.1) | 333 | 140 | 0.964 (0.942-0.980) | 93.4 (90.7-95.8) | 84.3 (77.9-90.0) |
| **Age** |  |  |  |  |  |  |  |  |  |  |
| ≤65 years | 196 | 102 | 0.980 (0.964-0.991) | 91.8 (88.3-95.4) | 89.2 (82.4-95.1) | 228 | 127 | 0.974 (0.958-0.986) | 93.4 (90.4-96.5) | 85.8 (79.5-92.1) |
| >65 years | 288 | 124 | 0.961 (0.941-0.978) | 94.1 (91.3-96.5) | 83.1 (76.6-89.5) | 363 | 166 | 0.960 (0.940-0.975) | 93.1 (90.4-95.6) | 84.9 (79.5-90.4) |
| **Ethnicity** |  |  |  |  |  |  |  |  |  |  |
| Hispanic | 41 | 18 | 1.000 (1.000-1.000) | 97.6 (92.7-100.0) | 100.0 (100.0-100.0) | 48 | 22 | 0.994 (0.974-1.000) | 91.7 (83.3-97.9) | 95.5 (86.4-100.0) |
| Not Hispanic | 411 | 196 | 0.969 (0.955-0.980) | 92.7 (90.0-95.1) | 86.2 (81.1-90.8) | 506 | 257 | 0.966 (0.953-0.978) | 93.3 (90.9-95.5) | 86.4 (81.7-90.3) |
| Prefer not to say / Decline | 2 | 1 | 1.000 (1.000-1.000) | 100.0 (100.0-100.0) | 100.0 (100.0-100.0) | 3 | 1 | 1.000 (1.000-1.000) | 100.0 (100.0-100.0) | 100.0 (100.0-100.0) |
| Unavailable | 30 | 11 | 0.897 (0.758-0.988) | 93.3 (83.3-100.0) | 54.5 (27.3-81.8) | 34 | 13 | 0.914 (0.814-0.980) | 94.1 (85.3-100.0) | 46.2 (23.1-69.2) |
| **Race** |  |  |  |  |  |  |  |  |  |  |
| American Indian or Alaska Native | 0 | 0 | - | - | - | 0 | 1 | - | - | 100.0 (100.0-100.0) |
| Asian | 35 | 11 | 0.971 (0.906-1.000) | 94.3 (85.7-100.0) | 72.7 (45.5-100.0) | 39 | 11 | 0.944 (0.853-0.995) | 92.3 (84.6-100.0) | 72.7 (45.5-100.0) |
| Black or African American | 30 | 12 | 0.978 (0.917-1.000) | 90.0 (76.7-100.0) | 91.7 (75.0-100.0) | 39 | 12 | 0.974 (0.915-1.000) | 94.9 (87.2-100.0) | 91.7 (75.0-100.0) |
| Native Hawaiian or Other Pacific Islander | 1 | 0 | - | 100.0 (100.0-100.0) | - | 2 | 0 | - | 100.0 (100.0-100.0) | - |
| White or Caucasian | 365 | 179 | 0.965 (0.948-0.979) | 93.2 (90.4-95.6) | 85.5 (79.9-90.5) | 455 | 239 | 0.964 (0.949-0.976) | 93.2 (90.8-95.4) | 85.8 (81.2-90.4) |
| Other | 29 | 11 | 1.000 (1.000-1.000) | 96.6 (89.7-100.0) | 100.0 (100.0-100.0) | 29 | 14 | 1.000 (1.000-1.000) | 93.1 (82.8-100.0) | 100.0 (100.0-100.0) |
| Two or more | 3 | 3 | 1.000 (1.000-1.000) | 66.7 (0.0-100.0) | 100.0 (100.0-100.0) | 4 | 4 | 1.000 (1.000-1.000) | 75.0 (25.0-100.0) | 100.0 (100.0-100.0) |
| Declined | 2 | 2 | 1.000 (1.000-1.000) | 100.0 (100.0-100.0) | 100.0 (100.0-100.0) | 2 | 2 | 1.000 (1.000-1.000) | 100.0 (100.0-100.0) | 100.0 (100.0-100.0) |
| Unavailable | 19 | 8 | 0.987 (0.921-1.000) | 94.7 (84.2-100.0) | 75.0 (50.0-100.0) | 21 | 10 | 0.967 (0.886-1.000) | 95.2 (85.7-100.0) | 50.0 (20.0-80.0) |
| **Manufacturer** |  |  |  |  |  |  |  |  |  |  |
| GE Healthcare | 107 | 59 | 0.968 (0.941-0.988) | 94.4 (89.7-98.1) | 84.7 (74.6-93.2) | 185 | 100 | 0.968 (0.949-0.983) | 93.5 (89.7-96.8) | 84.0 (76.0-91.0) |
| NeuroLogica | 1 | 0 | - | 100.0 (100.0-100.0) | - | 2 | 0 | - | 50.0 (0.0-100.0) | - |
| Philips | 1 | 0 | - | 100.0 (100.0-100.0) | - | 1 | 0 | - | 100.0 (100.0-100.0) | - |
| Siemens | 372 | 166 | 0.969 (0.953-0.982) | 92.7 (90.1-95.4) | 86.1 (80.7-91.0) | 368 | 162 | 0.967 (0.950-0.981) | 93.2 (90.5-95.7) | 86.4 (80.9-91.4) |
| Toshiba | 3 | 1 | 1.000 (1.000-1.000) | 100.0 (100.0-100.0) | 100.0 (100.0-100.0) | 35 | 31 | 0.973 (0.933-0.996) | 94.3 (85.7-100.0) | 83.9 (71.0-96.8) |

**Supplementary Table 10**: Number of cases for intraventricular hemorrhage cohort. *The number of selected cases was based on powering calculations; the ground truth radiologists did not have access to the original radiology reports and the interpretation could change. **One case was incorrectly processed during the series selection, image quality review and DICOM review when it should have been excluded; its later removal resulted in the cohort having one fewer case than initially planned and is reflected in from the second row of this table onwards (which has one fewer case than the first row).

|  | Positive cases | Negative cases | Total cases |
| --- | --- | --- | --- |
| **Original cases** |  |  |  |
| Number of cases selected based on original radiology reports* | 100 | 100 | 200 |
| Number of cases following ground truth radiologist interpretations** | 97 | 102 | 199 |
| **Thin series (≤1.5mm)** |  |  |  |
| Number of thin series | 79 | 80 | 159 |
| Number of thin series with successful model inference | 74 | 79 | 153 |
| **Thick series (>1.5mm and ≤5mm)** |  |  |  |
| Number of thick series | 97 | 102 | 199 |
| Number of thick series with successful model inference | 91 | 96 | 187 |

**Supplementary Table 11**: Model performance for identification of intraventricular hemorrhage at prespecified operating points.

| Operating point | True positive | False negative | Sensitivity (95% CI) | True negative | False positive | Specificity (95% CI) |
| --- | --- | --- | --- | --- | --- | --- |
| **Thin series** |  |  |  |  |  |  |
| 0.051859 | 67 | 7 | 90.5 (83.8-95.9) | 77 | 2 | 97.5 (93.7-100.0) |
| **Thick series** |  |  |  |  |  |  |
| 0.008430 | 87 | 4 | 95.6 (91.2-98.9) | 83 | 13 | 86.5 (79.2-92.7) |
| 0.015487 | 84 | 7 | 92.3 (86.8-96.7) | 86 | 10 | 89.6 (83.3-94.8) |
| 0.051859 | 80 | 11 | 87.9 (81.3-93.4) | 94 | 2 | 97.9 (94.8-100.0) |

**Supplementary Table 12**: Demographic and technical subgroup performance for identifying intraventricular hemorrhage.

|  | **Thin Series (Operating Point 0.051859)** | | | | | **Thick Series (Operating Point 0.051859)** | | | | |
| --- | --- | --- | --- | --- | --- | --- | --- | --- | --- | --- |
|  | Positive N | Negative N | AUC (95% CI) | Sensitivity (95% CI) | Specificity (95% CI) | Positive N | Negative N | AUC (95% CI) | Sensitivity (95% CI) | Specificity (95% CI) |
| **Overall** | 74 | 79 | 0.987 (0.969-0.997) | 90.5 (83.8-95.9) | 97.5 (93.7-100.0) | 91 | 96 | 0.983 (0.968-0.994) | 87.9 (81.3-93.4) | 97.9 (94.8-100.0) |
| **Sex** |  |  |  |  |  |  |  |  |  |  |
| Female | 40 | 37 | 1.000 (0.996-1.000) | 97.5 (92.5-100.0) | 100.0 (100.0-100.0) | 46 | 46 | 0.996 (0.986-1.000) | 91.3 (82.6-97.8) | 100.0 (100.0-100.0) |
| Male | 34 | 42 | 0.974 (0.936-0.996) | 82.4 (67.6-94.1) | 95.2 (88.1-100.0) | 45 | 50 | 0.973 (0.941-0.992) | 84.4 (73.3-93.4) | 96.0 (90.0-100.0) |
| **Age** |  |  |  |  |  |  |  |  |  |  |
| ≤65 years | 25 | 32 | 0.989 (0.955-1.000) | 88.0 (72.0-100.0) | 100.0 (100.0-100.0) | 30 | 41 | 0.991 (0.963-1.000) | 86.7 (73.3-96.7) | 100.0 (100.0-100.0) |
| >65 years | 49 | 47 | 0.985 (0.961-0.998) | 91.8 (83.7-98.0) | 95.7 (89.4-100.0) | 61 | 55 | 0.981 (0.959-0.994) | 88.5 (80.3-96.7) | 96.4 (90.9-100.0) |
| **Ethnicity** |  |  |  |  |  |  |  |  |  |  |
| Hispanic | 9 | 9 | 0.975 (0.852-1.000) | 88.9 (66.7-100.0) | 100.0 (100.0-100.0) | 9 | 9 | 1.000 (1.000-1.000) | 88.9 (66.7-100.0) | 100.0 (100.0-100.0) |
| Not Hispanic | 63 | 68 | 0.989 (0.972-0.998) | 90.5 (82.5-96.8) | 97.1 (92.6-100.0) | 78 | 85 | 0.984 (0.967-0.994) | 87.2 (79.5-93.6) | 97.6 (94.1-100.0) |
| Prefer not to say / Decline | 0 | 1 | - | - | 100.0 (100.0-100.0) | 0 | 1 | - | - | 100.0 (100.0-100.0) |
| Unavailable | 2 | 1 | 1.000 (1.000-1.000) | 100.0 (100.0-100.0) | 100.0 (100.0-100.0) | 4 | 1 | 1.000 (1.000-1.000) | 100.0 (100.0-100.0) | 100.0 (100.0-100.0) |
| **Race** |  |  |  |  |  |  |  |  |  |  |
| American Indian or Alaska Native | 0 | 0 | - | - | - | 0 | 0 | - | - | - |
| Asian | 2 | 2 | 1.000 (1.000-1.000) | 100.0 (100.0-100.0) | 100.0 (100.0-100.0) | 2 | 2 | 1.000 (1.000-1.000) | 100.0 (100.0-100.0) | 100.0 (100.0-100.0) |
| Black or African American | 7 | 9 | 1.000 (0.905-1.000) | 85.7 (57.1-100.0) | 100.0 (100.0-100.0) | 8 | 8 | 1.000 (0.937-1.000) | 87.5 (62.5-100.0) | 100.0 (100.0-100.0) |
| Native Hawaiian or Other Pacific Islander | 0 | 0 | - | - | - | 0 | 0 | - | - | - |
| White or Caucasian | 58 | 61 | 0.991 (0.975-1.000) | 91.4 (82.8-98.3) | 96.7 (91.8-100.0) | 72 | 80 | 0.985 (0.969-0.995) | 87.5 (79.2-94.4) | 97.5 (93.8-100.0) |
| Other | 5 | 4 | 0.900 (0.600-1.000) | 80.0 (40.0-100.0) | 100.0 (100.0-100.0) | 5 | 4 | 1.000 (1.000-1.000) | 80.0 (40.0-100.0) | 100.0 (100.0-100.0) |
| Two or more | 0 | 1 | - | - | 100.0 (100.0-100.0) | 0 | 0 | - | - | - |
| Declined | 0 | 1 | - | - | 100.0 (100.0-100.0) | 0 | 1 | - | - | 100.0 (100.0-100.0) |
| Unavailable | 2 | 1 | 1.000 (1.000-1.000) | 100.0 (100.0-100.0) | 100.0 (100.0-100.0) | 4 | 1 | 1.000 (1.000-1.000) | 100.0 (100.0-100.0) | 100.0 (100.0-100.0) |
| **Manufacturer** |  |  |  |  |  |  |  |  |  |  |
| GE Healthcare | 24 | 25 | 0.990 (0.960-1.000) | 91.7 (79.2-100.0) | 96.0 (88.0-100.0) | 24 | 24 | 0.997 (0.979-1.000) | 83.3 (66.7-95.8) | 100.0 (100.0-100.0) |
| NeuroLogica | 1 | 1 | 1.000 (1.000-1.000) | 0.0 (0.0-0.0) | 100.0 (100.0-100.0) | 1 | 1 | 1.000 (1.000-1.000) | 100.0 (100.0-100.0) | 100.0 (100.0-100.0) |
| Philips | 0 | 0 | - | - | - | 0 | 0 | - | - | - |
| Siemens | 49 | 53 | 0.987 (0.960-1.000) | 91.8 (83.7-98.0) | 98.1 (94.3-100.0) | 48 | 49 | 0.974 (0.940-0.994) | 85.4 (75.0-93.8) | 98.0 (93.9-100.0) |
| Toshiba | 0 | 0 | - | - | - | 18 | 22 | 0.995 (0.970-1.000) | 100.0 (100.0-100.0) | 95.5 (86.4-100.0) |
